# Acute Brain Ischemia, Infarction and Hemorrhage in Subjects Dying with or Without Autopsy-Proven Acute Pneumonia

**DOI:** 10.1101/2021.03.22.21254139

**Authors:** Thomas G. Beach, Lucia I. Sue, Anthony J. Intorcia, Michael J. Glass, Jessica E. Walker, Richard Arce, Courtney M. Nelson, Geidy E. Serrano

**Author notes:** Correspondence: Thomas G. Beach, Banner Sun Health Research Institute, 10515 West Santa Fe Drive, Sun City, AZ 85351.

## Abstract

Stroke is one of the most serious complications of Covid-19 disease but it is still unclear whether stroke is more common with Covid-19 pneumonia as compared to non-Covid-19 pneumonia. We investigated the concurrence rate of autopsy-confirmed acute brain ischemia, acute brain infarction and acute brain hemorrhage with autopsy-proven acute non-Covid pneumonia in consecutive autopsies in the Arizona Study of Aging and Neurodegenerative Disorders (AZSAND), a longitudinal clinicopathological study of normal aging and neurodegenerative diseases. Of 691 subjects with a mean age of 83.4 years, acute pneumonia was histopathologically diagnosed in 343 (49.6%); the concurrence rates for histopathologically-confirmed acute ischemia, acute infarction or subacute infarction was 14% and did not differ between pneumonia and non-pneumonia groups while the rates of acute brain hemorrhage were 1.4% and 2.0% of those with or without acute pneumonia, respectively. In comparison, in reviews of Covid-19 publications, reported clinically-determined rates of acute brain infarction range from 0.5% to 20% while rates of acute brain hemorrhage range from 0.13% to 2%. In reviews of Covid-19 autopsy studies, concurrence rates for both acute brain infarction and acute brain hemorrhage average about 10%. Covid-19 pneumonia and non-Covid-19 pneumonia may have similar risks tor concurrent acute brain infarction and acute brain hemorrhage when pneumonia is severe enough to cause death. Additionally, acute brain ischemia, infarction or hemorrhage may not be more common in subjects dying of acute pneumonia than in subjects dying without acute pneumonia.

## Introduction

Studies of factors associated with acute pneumonia, a leading cause of death worldwide ^1^, may have relevance for Covid-19 disease as both occur predominantly in the setting of epidemic viral respiratory infections ^2-4^. Predisposing factors to both acute non-Covid-19 pneumonia and epidemic Covid-19 disease include wintertime occurrence, older age, and male sex, while obesity, pre-existing cardiopulmonary conditions, and diabetes are particularly associated with Covid-19 disease ^5-26^. Also implicated as risk factors for both are age-related neurodegenerative diseases that cause parkinsonism and dementia ^27-44^.

Stroke, both ischemic and hemorrhagic, is a catastrophic accompaniment of Covid-19 disease but the concurrence rate is difficult to ascertain due to a very wide range in existing clinical and autopsy-based reports ^45-78^. Also, it is uncertain whether either Covid-19 disease or non-Covid-19 pneumonia are more likely to be associated with stroke, or whether stroke is more or less common in patients hospitalized for other reasons. As diagnosis of both pneumonia and stroke are more accurately obtained at autopsy ^79-83^ we investigated the concurrence of autopsy-proven pneumonia and acute stroke in the Arizona Study of Aging and Neurodegenerative Disorders (AZSAND), a longitudinal, clinicopathological study of normal aging and neurodegenerative diseases ^84^. Volunteer participants are primarily elderly residents of metropolitan Phoenix, Arizona. The program includes annual clinical assessments of community volunteers by neuropsychologists and subspecialty neurologists, with eventual whole-body autopsies by medically-certified anatomical pathologists and a neuropathologist. The current study was undertaken to determine the frequency of ischemic and hemorrhagic intracranial lesions in autopsied participants with or without autopsy-diagnosed acute pneumonia, prior to the onset of the Covid-19 pandemic.

We have previously reported on clinical and pathological associations of acute pneumonia in this same set of subjects, finding that significant risk factors of acute pneumonia include wintertime occurrence, older age, male sex and neurodegenerative dementia or parksinsonism ^27^.

## Methods

### Study subjects and clinical assessments

The study population draws primarily on the retirement communities of greater Phoenix, Arizona. The residents are predominantly elderly, well-educated, Caucasian (greater than 90%), middle- and upper-income individuals. Recruitment is directed at subjects with a clinical diagnosis of dementia, parkinsonism or cancer, or who are free of major neurological conditions. More than 70% of clinically-followed subjects are living independently at the time of enrollment, without cognitive impairment or parkinsonism. Subjects with hazardous infectious diseases including HIV, hepatitis B or C, Creutzfeldt-Jakob disease and other infectious encephalopathies are excluded from enrollment.

The AZSAND, through its Brain and Body Donation Program (https://www.brainandbodydonationregistration.org/) performs constitutively-rapid autopsies (median postmortem interval 3.0 hours for all 2,000 + autopsies) on Program participants and shares tissue samples with biomedical researchers worldwide ^84^. The Program is approved by an Institutional Review Board and written informed consent is obtained from all subjects or their legal representatives.

Standardized general medical, subspecialty neurological (behavioral/cognitive and movement disorders), and neuropsychological assessments are administered to most subjects annually. Clinical diagnostic classification is performed after each annual assessment, at a consensus conference attended by neurologists and neuropsychologists. A final clinicopathological diagnosis is assigned after death, after review of all standardized research clinical data, private medical records and neuropathological examination findings. The cause of death is recorded by the subjects’ physician without knowledge of the eventual autopsy findings.

### Autopsy examinations

Medically-certified pathologists perform all examinations. Lung sampling is directed at both grossly-normal and grossly-pathological regions. Acute pneumonia is diagnosed on the basis of microscopic findings, in hematoxylin and eosin-stained slides, of confluent intra-alveolar accumulations of neutrophil polymorphonuclear leukocytes, with or without a typical gross pathological appearance or microscopic evidence of bacterial colonies. The neuropathological diagnostic approach has been previously described and is supplemented by consideration of results of standardized annual neurological and neuropsychological assessments ^84^. Published clinicopathological consensus criteria ^85-92^ are used when they exist, incorporating clinical assessment results as well as pertinent private medical history. The presence of absence in the brain of acute or subacute infarcts, as well as acute hemorrhage, is recorded. Infarcts are subclassified as large (> 27 cc), small (2-27 cc), lacunar (grossly visible, 1-2 cc or less) or microscopic (not grossly visible, less than 1 cc). Acute ischemic changes without frank infarction are also recorded, including eosinophilic perikaryal neuronal cytoplasm, nuclear pyknosis, karyorrhexis or dissolution, loss of tissue eosinophilia and tissue microvacuolation. Hemorrhage is classified as large (> 27 cc) or small/microscopic.

### Associations tested and statistical methods

Fisher’s Exact Tests were used to test for associations of subject characteristics with pneumonia and different types of stroke or acute ischemic changes.

## Results

Included in this study were 691 subjects with postmortem lung and brain examinations, done between March 2005 and December 2019 (Table 1). Of these, 405 were males (59%) and 286 were females (41%) with mean ages of 82.1 and 85.2 years, respectively. Acute pneumonia was diagnosed histopathologically in 343 subjects (49.6%; 225 males and 118 females). This was almost entirely acute bronchopneumonia with a very small percentage of acute lobar pneumonia. In contrast, pneumonia or respiratory failure was listed as the cause of death (by the physician on the basis of clinical findings) in less than 1% of cases.

**Table 1.** Neuropathological and clinical classification of study subjects. Norm = non-demented without parkinsonism; AD = Alzheimer’s disease; PD = Parkinson’s disease; DLB = dementia with Lewy bodies; VaD = vascular dementia; PSP = progressive supranuclear palsy.

|  | **ALL** | **Norm** | **AD** | **PD** | **DLB** | **VaD** | **PSP** |
| --- | --- | --- | --- | --- | --- | --- | --- |
| **Number** | 691 | 185 | 319 | 127 | 72 | 78 | 49 |
| **Age (SD)** | 83.4 (9.6) | 85 (3.5) | 84.3 (2.8) | 80 (2.1) | 83.6 (9.2) | 89.3 (10.6) | 86.4 (6.8) |
| **M/F** | 405/286 | 112/73 | 182/137 | 89/38 | 51/21 | 39/39 | 30/19 |

Histopathologically-confirmed acute ischemia, acute infarction or subacute infarction was present in 14% of brains from autopsied subjects, from both the pneumonia and non-pneumonia groups, while acute brain hemorrhage was present in 1.4% and 2.0% of those with or without acute pneumonia, respectively (Table 2). Of subtypes of infarction or ischemic changes and hemorrhage, only acute ischemic changes were significantly more common with pneumonia (2.6% vs 0.57%; p = 0.036%). Small or microscopic hemorrhages were present in 4 pneumonia cases but were not present in any non-pneumonia case. Stroke was listed as the clinical cause of death in less than 6% of cases.

**Table 2.**
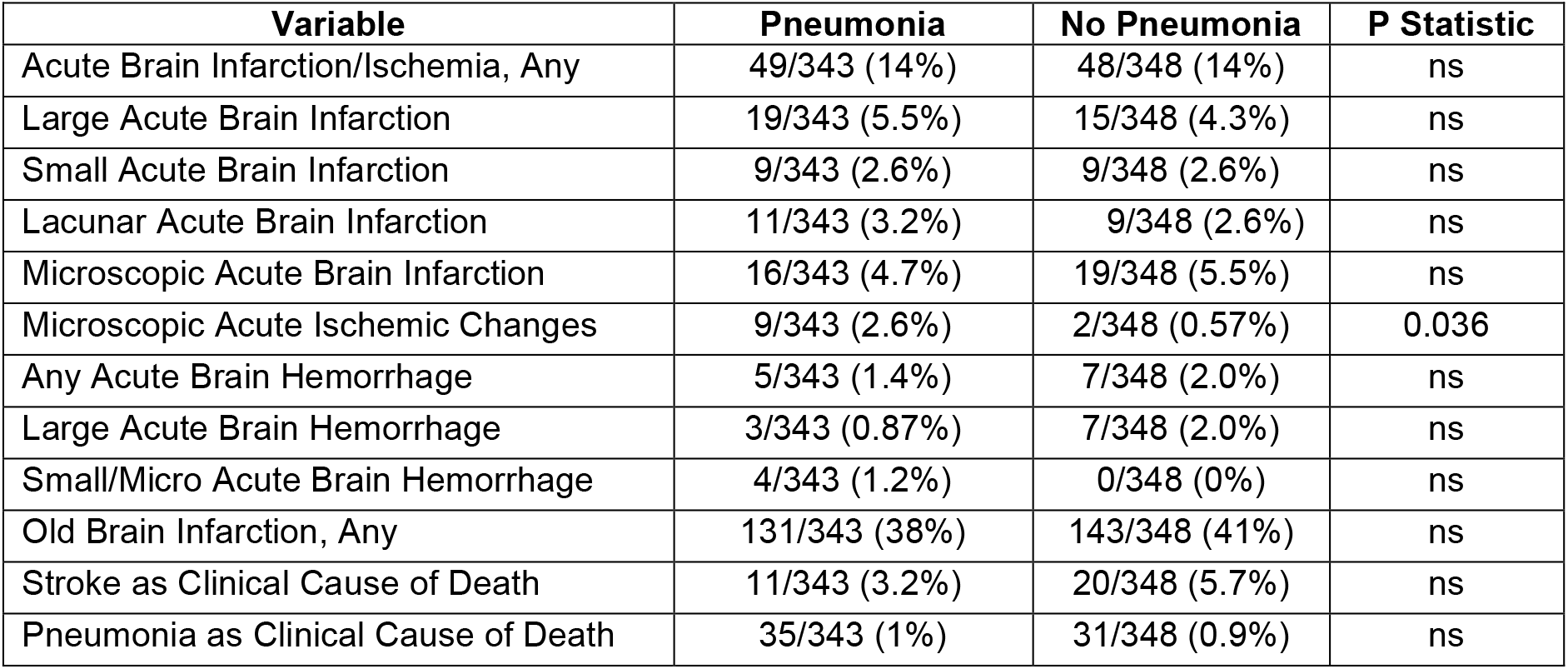
Concurrence rates of histopathological evidence of acute or subacute brain infarction or ischemia or acute hemorrhage, or of acute stroke as a listed clinical cause of death, in subjects with and without a histopathological diagnosis of acute pneumonia. Fisher Exact tests were used to compare proportions.

| **Variable** | **Pneumonia** | **No Pneumonia** | **P Statistic** |
| --- | --- | --- | --- |
| Acute Brain Infarction/Ischemia, Any | 49/343 (14%) | 48/348 (14%) | ns |
| Large Acute Brain Infarction | 19/343 (5.5%) | 15/348 (4.3%) | ns |
| Small Acute Brain Infarction | 9/343 (2.6%) | 9/348 (2.6%) | ns |
| Lacunar Acute Brain Infarction | 11/343 (3.2%) | 9/348 (2.6%) | ns |
| Microscopic Acute Brain Infarction | 16/343 (4.7%) | 19/348 (5.5%) | ns |
| Microscopic Acute Ischemic Changes | 9/343 (2.6%) | 2/348 (0.57%) | 0.036 |
| Any Acute Brain Hemorrhage | 5/343 (1.4%) | 7/348 (2.0%) | ns |
| Large Acute Brain Hemorrhage | 3/343 (0.87%) | 7/348 (2.0%) | ns |
| Small/Micro Acute Brain Hemorrhage | 4/343 (1.2%) | 0/348 (0%) | ns |
| Old Brain Infarction, Any | 131/343 (38%) | 143/348 (41%) | ns |
| Stroke as Clinical Cause of Death | 11/343 (3.2%) | 20/348 (5.7%) | ns |
| Pneumonia as Clinical Cause of Death | 35/343 (1%) | 31/348 (0.9%) | ns |

Clinicopathological brain diagnoses other than stroke were predominantly a variety of neurodegenerative diseases, including 319 dementia subjects with Alzheimer’s disease (AD), 127 subjects with idiopathic Parkinson’s disease (PD), 72 subjects with dementia with Lewy bodies (DLB), 78 with vascular dementia (VaD) and 49 with diagnostic histopathology of progressive supranuclear palsy (PSP). These diagnoses are not mutually exclusive as many of the subjects had more than one neuropathological diagnosis. Additionally, 185 subjects had neither dementia or parkinsonism and had less-than-diagnostic neurodegenerative changes. Of the 319 AD cases, 34 also had clinicopathologically-diagnosed PD, 62 had DLB, 60 had VaD and 16 had progressive supranuclear palsy. Other neurodegenerative conditions diagnosed but not statistically tested for associations with pneumonia or stroke due to low subject numbers included frontotemporal lobar degeneration with TDP-43 proteinopathy (n = 14), corticobasal degeneration (n = 7) and Pick’s disease (n = 4.)

A neuropathological diagnosis of vascular dementia and a clinical diagnosis of atrial fibrillation were both significantly more likely to have had an autopsy diagnosis of acute brain ischemia or infarction (Table 3). Female subjects, subjects over age 85, subjects with severe circle of Willis atherosclerosis and congestive heart failure were more likely, and subjects with a neuropathological diagnosis of AD were less likely to have had acute brain ischemia or infarction; these associations did not reach significance levels.

**Table 3.** Comparison of subject characteristics of those with and without histopathologically-proven acute or subacute brain ischemia or infarction. Fisher Exact tests were used to compare proportions. Obesity is defined as BMI 30 or greater. Cardiomegaly is defined as heart weight > 360 g for males and > 280 g for females. Older age is defined as greater than the median age of 84. Cerebral atherosclerosis is defined as an autopsy-based circle of Willis score of severe. Any ApoE4 = apolipoprotein E genotype heterozygous or homozygous for the ε4 allele.

| Variable | Acute Ischemia/Infarction | No Acute Ischemia/Infarction | P Statistic |
| --- | --- | --- | --- |
| Stroke as Clinical Cause of Death | 16/343 (4.7%) | 15/348 (4.3%) | ns |
| Male | 53/97 (55%) | 325/594 (55%) | ns |
| Female | 44/97 (45%) | 242/594 (41%) | ns |
| Older Age | 56/97 (58%) | 280/594 (47%) | ns |
| AD | 35/97 (36%) | 284/594 (48%) | ns |
| PD | 17/97 (17.5%) | 110/594 (18.5%) | ns |
| DLB | 9/97 (9%) | 63/594 (6%) | ns |
| VaD | 21/97 (22%) | 56/594 (9%) | 0.0013 |
| PSP | 5/97 (5%) | 44/594 (7%) | ns |
| Any ApoE4<sup>1</sup> | 27/97 (28%) | 187/586 (32%) | ns |
| Non-Summer Death | 77/97 (79%) | 455/594 (77%) | ns |
| COPD | 25/97 (26%) | 160/594 (27%) | ns |
| Old brain infarction, Any | 43/97 (44%) | 231/594 (39%) | ns |
| Obesity (BMI 30 or greater)<sup>2</sup> | 26/97 (27%) | 153/571 (24%) | ns |
| Cardiomegaly<sup>3</sup> | 72/97 (74%) | 443/589 (75%) | ns |
| Circle of Willis atherosclerosis | 34/97 (35%) | 168/594 (28%) | ns |
| Atrial fibrillation | 37/97 (38%) | 159/594 (27%) | 0.03 |
| Congestive Heart Failure | 31/97 (32%) | 137/594 (23%) | ns |
| Hypertension | 73/97 (75%) | 421/594 (71%) | ns |
| Diabetes | 27/97 (28%) | 137/594 (23%) | ns |
| Cigarette Smoker | 37/97 (38%) | 277/594 (47%) | ns |

1. Eight subjects without acute ischemia/infarction had no apolipoprotein E genotyping available. 2. Twenty-three subjects without acute ischemia/infarction had no BMI values available. 3. Five subjects without acute ischemia/infarction had no heart weight available.

## Discussion

Although stroke and a wide range of neurological signs and symptoms have been reported as relatively common accompaniments of Covid-19 disease, most reports have lacked an appropriate control group. As direct CNS infection with SARS-CoV-2 may occur only in a minority of those with Covid-19 disease ^93^, much of the reported neurological phenomena may occur not through direct SARS-CoV-2 infection of neural tissue but indirectly, via systemic complications of critical illness such as sepsis, coagulopathy, hyperimmune reactions or multi-organ failure. Better estimates of the specificity of any non-pulmonary Covid-19 accompaniments will only be possible by comparison with their concurrence rates in other types of critical illness.

Stroke is perhaps the most serious non-pulmonary complication of Covid-19 but its concurrent prevalence is difficult to ascertain due to a very wide range in existing clinical and autopsy-based reports ^45-78^. Non-Covid-19 pneumonia may be a useful control condition with which to approach this issue, as like Covid-19 disease, it is a life-threatening lung infection that frequently has systemic effects including sepsis, coagulopathy and multi-organ failure ^94^. As the diagnosis of both pneumonia and stroke are more accurately obtained at autopsy ^79-83^ we investigated the concurrence of autopsy-proven pneumonia and acute stroke in the Arizona Study of Aging and Neurodegenerative Disorders (AZSAND), a longitudinal, clinicopathological study of normal aging and neurodegenerative diseases ^84^.

In this study of a suburban-dwelling, predominantly Caucasian middle-class population of advanced age, acute ischemic changes and acute infarctions were common autopsy findings, occurring in 14% of subjects regardless of the presence or absence of concurrent acute pneumonia. This finding supports previous studies that indicate that acute ischemia and acute infarction are very common in subjects with critical illnesses resulting in death, regardless of whether pneumonia is also present, and that these strokes are often clinically unrecognized ^95^. The rates of clinical cause-of-death attribution, for both pneumonia and stroke in our study, are even lower than for reported clinicopathological studies; this is likely because the majority of subjects in our program die on hospice care and thus do not receive elaborate perimortem medical investigation or treatment.

Acute brain hemorrhage was an infrequent finding, with insufficient case numbers for statistical evaluation. We found significant associations of acute brain ischemia or infarction with a history of atrial fibrillation, as well as a diagnosis of vascular dementia, both of which are well known to be associated with higher risks of stroke ^96-100^.

In comparison, in reviews of Covid-19 publications, reported clinically and autopsy-determined rates of acute brain infarction range from 0.5% to 20% while rates of acute brain hemorrhage range from 0.13% to 9.5% ^45-78^. A small number of clinical studies have also compared rates of stroke in hospitalized patients with Covid-19 versus influenza, with two studies finding increased rates with Covid-19 ^74,75^ (1.6% vs 0.2% and 0.9% vs 0.3%) and one finding equivalent rates ^45^ T(1.2% for both). As these rates are all much smaller than what we have found here and as clinical detection of both stroke and pneumonia is poorly sensitive compared to autopsy ^79-83,95^, the overall likelihood is that Covid-19 pneumonia and non-Covid-19 pneumonia may have similar risks tor concurrent acute brain infarction and acute brain hemorrhage when pneumonia is severe enough to cause death, and stroke overall may not be differentially prevalent in subjects dying with either type of pneumonia or non-Covid-19 pneumonia as compared to subjects dying from other causes.

A limitation of our study is the lack of identification of the responsible pathogens. This is a common limitation to pneumonia studies ^101^. When pathogens are identified, they are viral more often than bacterial, with the top three in US hospitalized patients being human rhinovirus, influenza A and B, and Streptococcus pneumoniae ^102^. Although we did not attempt to determine, from clinical records or postmortem culture, the presence of associated pathogens, the confluent intra-alveolar accumulation of neutrophils is generally regarded as indicative of a bacterial cause ^103^ and therefore this study is likely to have been biased towards the inclusion of bacterial pneumonia and against the inclusion of purely viral pneumonia, although mixed bacterial and viral causes were very likely to have been common ^101 102 103^.

## Data Availability

All data is available on request at https://www.brainandbodydonationregistration.org/

## Acknowledgements

This project was supported by a Covid-19-focused supplement to a National Institute on Aging grant (3P30AG019610-20S1), submitted in response to a Notice of Special Interest (NOSI) issued by the National Institute on Aging (NOT-AG-20-022), “to highlight the urgent need for research on Coronavirus Disease 2019…”. The Brain and Body Donation Program has been supported by the National Institute of Neurological Disorders and Stroke (U24 NS072026 National Brain and Tissue Resource for Parkinson’s Disease and Related Disorders), the National Institute on Aging (P30 AG19610 Arizona Alzheimer’s Disease Core Center), the Arizona Department of Health Services (contract 211002, Arizona Alzheimer’s Research Center), the Arizona Biomedical Research Commission (contracts 4001, 0011, 05-901 and 1001 to the Arizona Parkinson’s Disease Consortium) and the Michael J. Fox Foundation for Parkinson’s Research.

